# Development and Multicenter Performance Evaluation of The First Fully Automated SARS-CoV-2 IgM and IgG Immunoassays

**DOI:** 10.1101/2020.04.16.20067231

**Authors:** Chungen Qian, Mi Zhou, Fangming Cheng, Xiaotao Lin, Yijun Gong, Xiaobing Xie, Ping Li, Zhiyong Li, Pingan Zhang, Zejin Liu, Fang Hu, Yun Wang, Quan Li, Yan Zhu, Guikai Duan, Yinting Xing, Huanyu Song, Wenfang Xu, Bi-Feng Liu, Fuzhen Xia

**Author notes:** Address correspondence to: F.X. at Shenzhen YHLO Biotech Co., Ltd, YHLO Bioscience Building, Baolong 2nd Rd, Longgang District, Shenzhen 518116, China. B.F.L. at College of Life Science and Technology, Huazhong University of Science and Technology, Wuhan 430074, China. These two authors contributed equally to this work (Chungen Qian, Mi Zhou).

## Abstract

**BACKGROUND:** The outbreak of severe acute respiratory syndrome coronavirus 2 (SARS-CoV-2) has rapidly spread globally. The laboratory diagnosis of SARS-CoV-2 infection has relied on nucleic acid tests. However, there are many limitations of nucleic acid tests, including low throughput and high rates of false negatives. More sensitive and accurate tests to effectively identify infected patients are needed.

**METHODS:** This study has developed fully automated chemiluminescent immunoassays (CLIA) to determine IgM and IgG antibodies to SARS-CoV-2 in human serum. The assay performance has been evaluated at 10 hospitals. Clinical specificity was evaluated by measuring 972 hospitalized patients with diseases other than COVID-19, and 586 donors of a normal population. Clinical sensitivity was assessed on 503 confirmed cases of SARS-CoV-2 by RT-PCR and 52 suspected cases.

**RESULTS:** The assays demonstrated satisfied assay precision with coefficient of variation (CV) of less than 4.45%. Inactivation of specimen does not affect assay measurement. SARS-CoV-2 IgM shows clinical specificity of 97.33% and 99.49% for hospitalized patients and normal population respectively. SARS-CoV-2 IgG shows clinical specificity of 97.43% and 99.15% for the hospitalized patients and the normal population respectively. SARS-CoV-2 IgM and IgG show clinical sensitivity of 85.88% and 96.62% respectively for confirmed SARS-Cov-2 infection with RT-PCR, of 73.08% and 86.54% respectively for suspected cases.

**CONCLUSIONS:** we have developed fully automated immunoassays for detecting SARS-CoV-2 IgM and IgG antibodies in human serum. The assays demonstrated high clinical specificity and sensitivity, and add great value to nucleic acid testing in fighting against the global pandemic of the SARS-CoV-2 infection.

## Introduction

Severe acute respiratory syndrome coronavirus 2 (SARS-CoV-2) was initially identified in Wuhan, Hubei province, China, in December 2019, causing an ongoing outbreak of the novel coronavirus disease (COVID-19) *(1,2)*. Although the epidemic in China has come under control through strict containment precautions over the past two months, SARS-CoV-2 has rapidly spread to more than 200 countries and regions *(3)*. SARS-CoV-2 belongs to the same family of coronaviruses as severe acute respiratory syndrome (SARS-CoV) and Middle East respiratory syndrome (MERS-CoV), however, its transmission efficiency is much higher than SARS-CoV and MERS-CoV, and there is no indication that transmission will cease as the weather gets warmer. The World Health Organization (WHO) has announced COVID-19 is now a global pandemic, and that there is a long battle ahead to fight the virus *(4)*.

At present, there is no effective medication to treat the disease and it might take a year for a vaccine to be developed. The only way to control the outbreak of the virus is to identify and quarantine the infected individuals. COVID-19 is characterized by similar symptoms as the common cold such as fever, non-productive cough, fatigue, and featured chest CT patterns that may also cause fatal complications, as with severe acute respiratory syndrome *(5, 6)*. The etiological diagnosis of SARS-CoV-2 infection relies on a nucleic acid test with Real Time-PCR (RT-PCR) using specimens collected via nasopharyngeal swab *(7)*. However, high numbers of false negatives have been found in some laboratories, leading to a positive detection rate of RT-PCR around 50% of suspected clinical and epidemiological COVID-19 cases. There are many factors that can cause false negatives, and the specimen collection via nasopharyngeal swab could be the major challenge as the chance for the virus to move to the upper respiratory tract is smaller via non-productive cough. Degradation of the virus mRNA due to specimen inactivation at 56°C is another cause for false negative result *(8)*.

The limited detection efficacy of RT-PCR for identifying SARS-CoV-2 infection requires the need for a more sensitive test to effectively identify COVID-19 patients for a better control of the virus transmission and proper treatment of the disease. Detection of specific serum antibodies for SARS-CoV-2 could provide an alternative solution and compensate for the limitations of the nucleic acid test. It has been well established that IgM antibodies are generated and released into the blood soon after the viral infection, followed by IgG antibodies, therefore the detection of the specific antibodies in blood is a sensitive measurement for viral infection. It has been reported that the IgM antibodies can be detected in the blood from 7 days, and begins to decrease 10 days after the disease onset, while IgG antibodies can be detected 8-9 days after the disease onset and continues to increase rapidly thereafter *(9)*. Commercial and non-commercial serological tests are currently under development.

Based on the sequence of SARS-CoV-2 nucleic acid, we have constructed and expressed the viral nucleocapsid protein and spike protein. Using the purified recombinant antigen, we developed chemiluminescent immunoassays (CLIA) to determine the IgM and IgG antibodies to SARS-CoV-2 in human serum or plasma, which is performed automatically by an immunoassay analyzer. The performance of the assays has been evaluated at 10 hospitals, demonstrating high specificity and high sensitivity for SARS-CoV-2 detection. Being an effective way to compensate for the false negative issue of RT-PCR, the fully automated SARS-CoV-2 IgM and IgG immunoassays provided a powerful means of laboratory diagnosis for fighting against the global spread of the virus.

## Materials and Methods

### PREPARATION OF RECOMBINANT PROTEINS OF SARS-COV-2

Using pFastBac1 vector, the plasmids were constructed by inserting gene fragments for expressing the SARS-CoV-2 nucleocapsid protein and spike protein based on the published SARS-CoV-2 nucleic acid sequence on Genbank (MN908947.3). They were then transfected into the insect cell Sf9 to express SARS-CoV-2 fusion proteins, which were then purified by a combination of affinity chromatography and ion-exchange chromatography. Screening and verification for the specificity and sensitivity of the recombined SARS-CoV-2 fusion protein were performed using SARS-CoV-2 IgM and IgG immunoassay system.

### DEVELOPMENT OF AUTOMATED CLIA FOR SARS-COV-2 IGM AND IGG

Paramagnetic carboxylated-microparticles (purchased from Thermo Scientific) were coated with the recombinant protein of SARS-CoV-2 through cross-linking by N-Ethyl-N’-(3-dimethylaminopropyl) carbodiimide (purchased from Thermo Scientific). Mouse monoclonal anti-human IgM or IgG (purchased from Fapon Biotech) were conjugated with NSP-DMAE-NHS (purchased from Maxchemtech), and the conjugated antibodies were then purified by gel filtration on a Sephadex G-50 column. The calibrators were made from inactivated SARS-CoV-2 human serum with designated concentrations in arbitrary unit (AU/mL). The Pre-Trigger and Trigger solutions are composed of sodium hydroxide and hydrogen peroxide. All measuring procedures were performed by the fully automated immune analyzer (manufactured by Shenzhen YHLO Biotech). The correlation between the chemiluminescent signal measured as relative light unit (RLU) and the concentration of SARS-CoV-2 IgM or IgG is shown in the dose-response curves (Figure 1).

**Figure 1:**
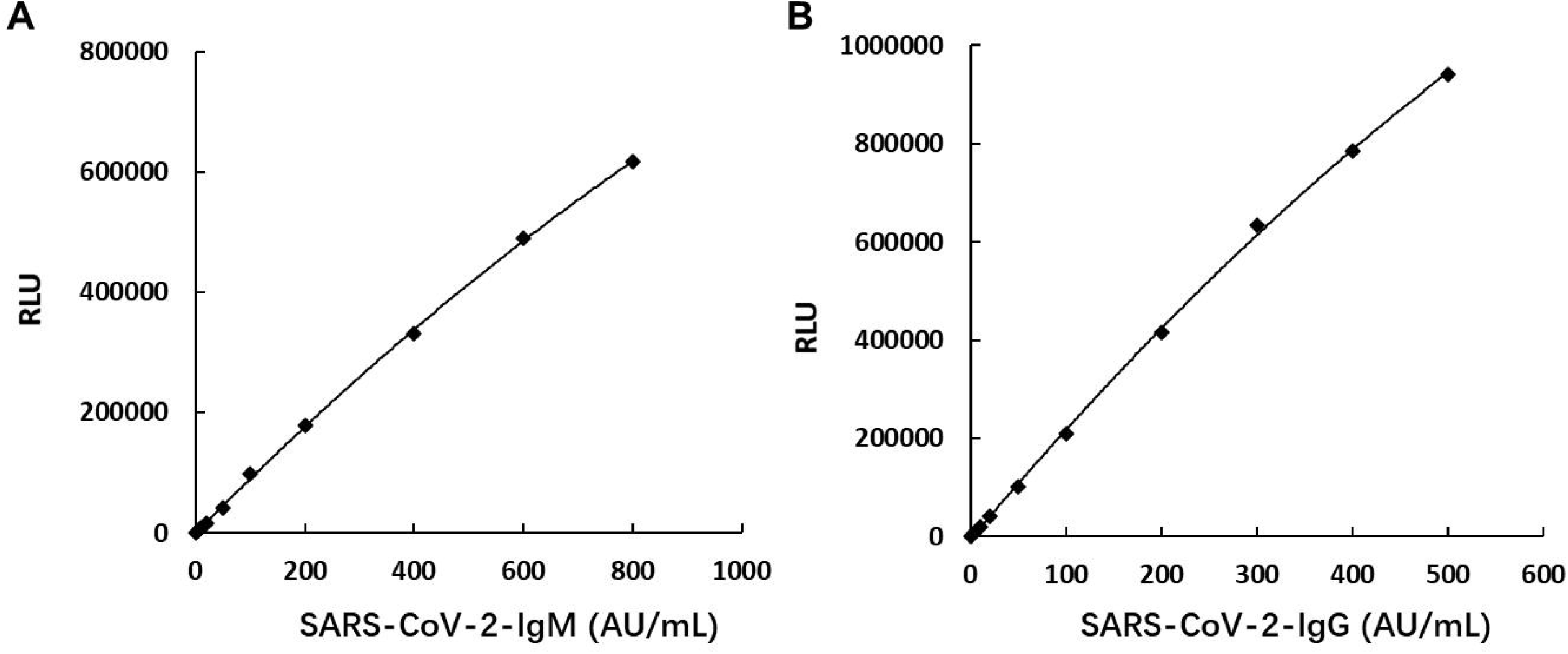
Dose-response curve of SARS-CoV-2 IgM (A) and SARS-CoV-2 IgG (B). The y-axis labelled “RLU” denotes the chemiluminescent signal measured as relative light unit.

### PRINCIPLE OF THE ASSAY

The SARS-CoV-2 IgM and IgG assays are two-step immunoassays for the qualitative detection of SARS-CoV-2 IgM and IgG antibodies in human serum and plasma, using direct chemiluminometric microparticle technology *(10, 11)*. The assays are performed on a fully automated immunoassay analyzer (manufactured by YHLO Biotech). In the first step, sample, recombinant SARS-Cov-2 antigens-coated paramagnetic microparticles, and a sample treating agent are combined. SARS-CoV-2 IgM or IgG antibodies present in the sample binds to the SARS-CoV-2 antigens-coated microparticles. After washing, acridinium-labeled anti-human IgM or IgG conjugate is added to form a reaction complex in the second step. Following another wash cycle, Pre-Trigger and Trigger solutions are added to the reaction mixture. The resulting chemiluminescent reaction is measured as RLUs. A direct relationship exists between the amount of SARS-CoV-2 IgM or IgG antibodies in the sample and the RLUs detected by the optical system of the immune analyzer. The concentration of SARS-CoV-2 IgM or IgG in the sample is determined by comparing the RLU of a sample to the RLU determined from two calibrators. If the concentration of the sample is greater than or equal to 10 AU/mL, the sample is considered reactive to SARS-CoV-2 IgM or IgG.

### PATIENT SAMPLES

The patient samples were collected from individuals recruited from 10 hospitals, 4 of which were from the worst outbreak in Hubei province, and the rest from 6 other provinces in China. The patients diagnosed with SARS-CoV-2 infection were confirmed by a RT-PCR nucleic acid test, and the suspected cases were assessed following guidelines of diagnosis and treatment of COVID-19, including typical epidemiological history, clinical symptoms and featured chest CT image *(12)*. The control subjects included hospitalized patients with diseases other than COVID-19, and donors of a normal population undergoing physical examinations. The study was conducted with the approval of the review board for human studies from each hospital attended.

### PERFORMANCE EVALUATION

Repeatability and within-laboratory precision were evaluated according to the Clinical and Laboratory Standards Institute (CLSI) EP5-A2 protocol *(13)*. One negative and two to three positive serums for SARS-CoV-2 IgM or IgG were used for the study. Each sample was measured in duplicate, two runs (morning and afternoon) per day over 20 testing days (n = 80). Repeatability and within-laboratory precision were calculated taking repeatability, run-to-run and day-to-day variance into account. Linearity was assessed according to CLSI EP6-A guidelines *(14)*. A sample with high SARS-CoV-2 IgM or IgG concentration was mixed in different proportions with a sample of low SARS-CoV-2 IgM or IgG concentration to form a dilution series. Each dilution was subsequently assayed in triplicates in one run, and mean results of the measured values were plotted against the dilution ratios. Serum samples negative and positive to SARS-CoV-2 IgM or IgG from 55 subjects (15 negative and 40 positive) were measured in duplicate before and after inactivation at 56°C for 30 minutes. Clinical specificity was evaluated by measuring 972 hospitalized patients with diseases other than COVID-19, and 586 donors of normal population undergoing physical examinations. Clinical sensitivity was assessed on patients diagnosed to SARS-CoV-2 infection either by a RT-PCR nucleic acid test (confirmed cases of 503 patients), or by typical epidemiological history, clinical symptoms and featured chest CT images (suspected cases of 52 patients).

### STATISTICAL ANALYSIS

For the precision study, a percentage of coefficient of variance (CV) of less than 20% was considered acceptable. For linearity study, the measured values were plotted against the dilution ratio and a regression model was selected according to CLSI EP6-A. For sample stability study, correlation and comparison of SARS-CoV-2 IgM or IgG values before and after inactivation were assessed by Passing-Bablok regression analysis and Bland-Altman difference plots. Clinical specificity and sensitivity were calculated according to the following formulas: Clinical specificity (%) = 100 × [True negative / (True Negative + False Positive)]; Clinical sensitivity (%) = 100 × [True Positive/ (True Positive + False Negative)].

## Results

### REPEATABILITY

Four serum samples (one negative and three positives for SARS-CoV-2 IgM) and three serum samples (one negative and two positives for SARS-CoV-2 IgG) were measured twice-a-day in duplicates over the span of 20 days, and 80 data points were obtained on each serum. Results are shown in Table 1 and Table 2. The repeatability of the SARS-CoV-2 IgM is from 2.80% to 4.32%, and the within-laboratory precision is from 3.02% to 4.45%. The repeatability of the SARS-CoV-2 IgG is from 3.11% to 4.30%, and the within-laboratory precision is from 3.12% to 5.13%.

**Table 1:** Repeatability and within-laboratory precision of SARS-CoV-2 IgM

| Sample | Mean<br>(AU/mL) | Repeatability |  | Within-Laboratory |  |
| --- | --- | --- | --- | --- | --- |
|  |  | SD (AU/mL) | CV (%) | SD (AU/mL) | CV (%) |
| Serum 1 | 7.29 | 0.31 | 4.32% | 0.32 | 4.45% |
| Serum 2 | 13.5 | 0.41 | 3.06% | 0.44 | 3.26% |
| Serum 3 | 98.2 | 2.75 | 2.80% | 2.97 | 3.02% |
| Serum 4 | 295 | 8.93 | 3.03% | 9.07 | 3.08% |

**Table 2:**
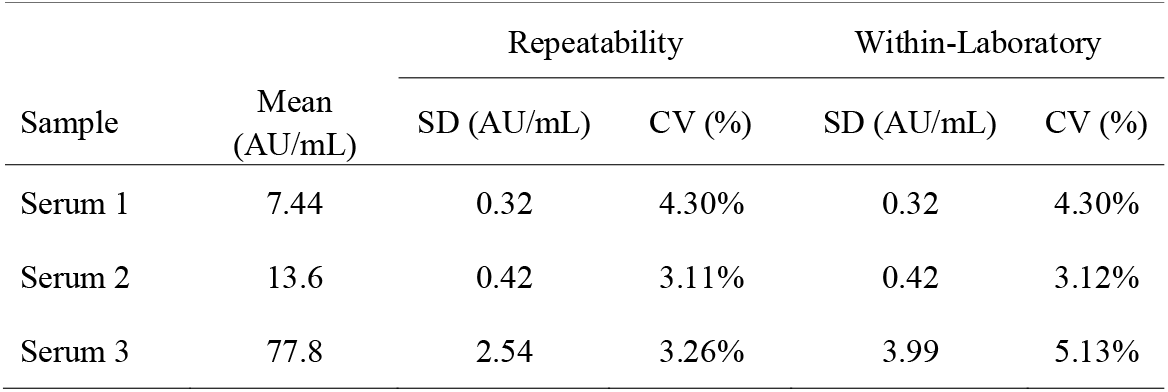
Repeatability and within-laboratory precision of SARS-CoV-2 IgG

### SAMPLE STABILITY AFTER INACTIVATION AT 56°C

Passing-Bablok regression and Bland-Altman analysis are shown in Figure 2. Across the concentration range of 55 serum samples, the concentrations of SARS-CoV-2 IgM and IgG measured after sample inactivation strongly correlate with that before sample inactivation. Spearman’s correlation coefficient is 0.993 for SARS-CoV-2 IgM, and 0.990 for SARS-CoV-2 IgG (p< 0.001 for both assays). No significant change of SARS-CoV-2 IgM and IgG concentration were observed after inactivation at 56°C for 30 minutes based on the Bland-Altman difference plots. Therefore, there is no worry to inactivate serum samples before measurement with SARS-CoV-2 IgM and IgG assays.

**Figure 2:**
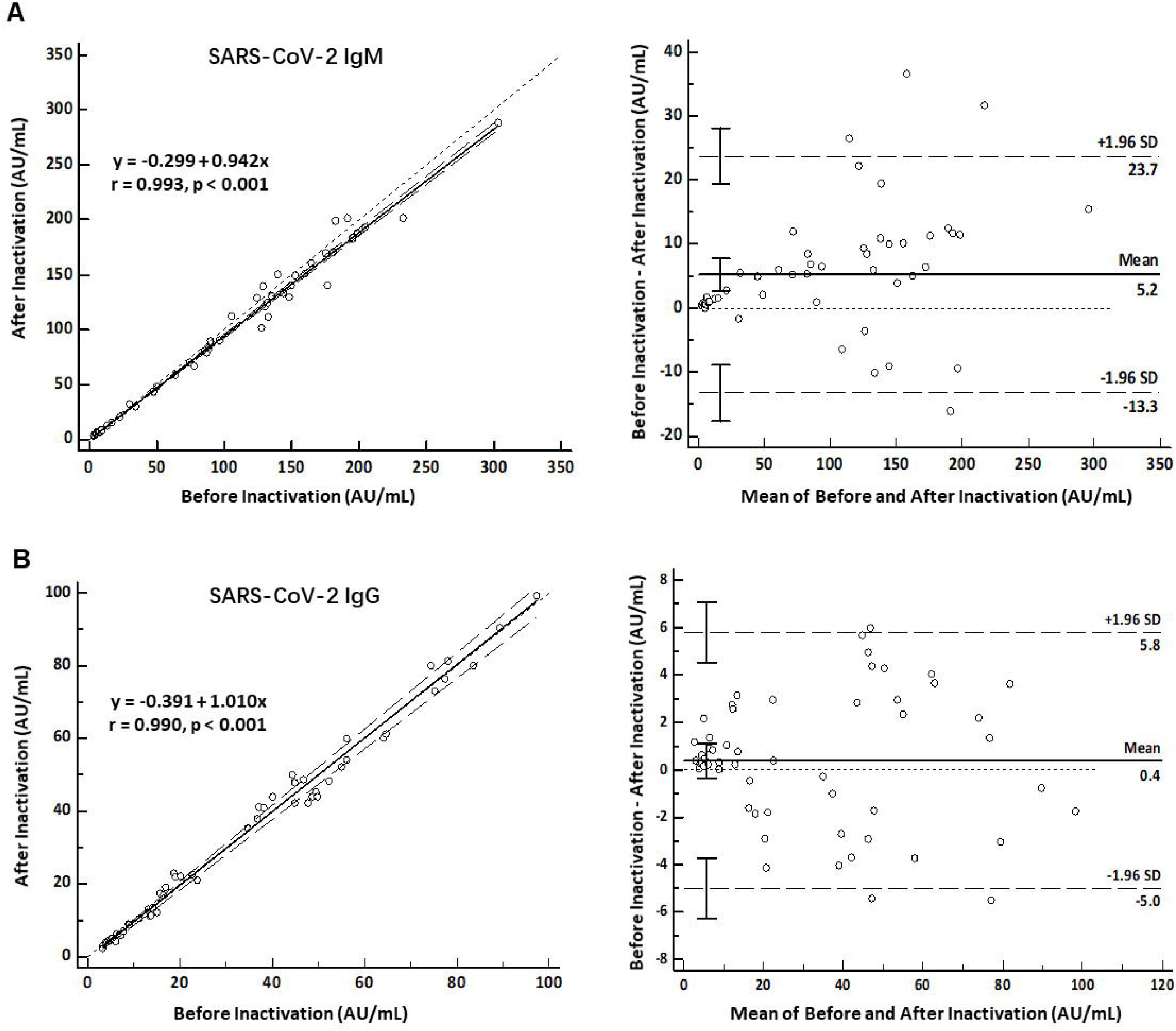
Passing-Bablok regression analysis (left panel) and Bland-Altman difference plots (right panel) for SARS-CoV-2 IgM and IgG concentrations measured before and after sample inactivation.

### LINEARITY

SARS-CoV-2 IgM assay shows good linearity (R^2^ = 0.9952) at the measuring range of 0.20 – 879.74 AU/mL (Figure 3A), and SARS-CoV-2 IgG assay also shows good linearity (R^2^ = 0.9982) at the measuring range of 0.20 – 453.50 AU/mL (Figure 3B)

**Figure 3:**
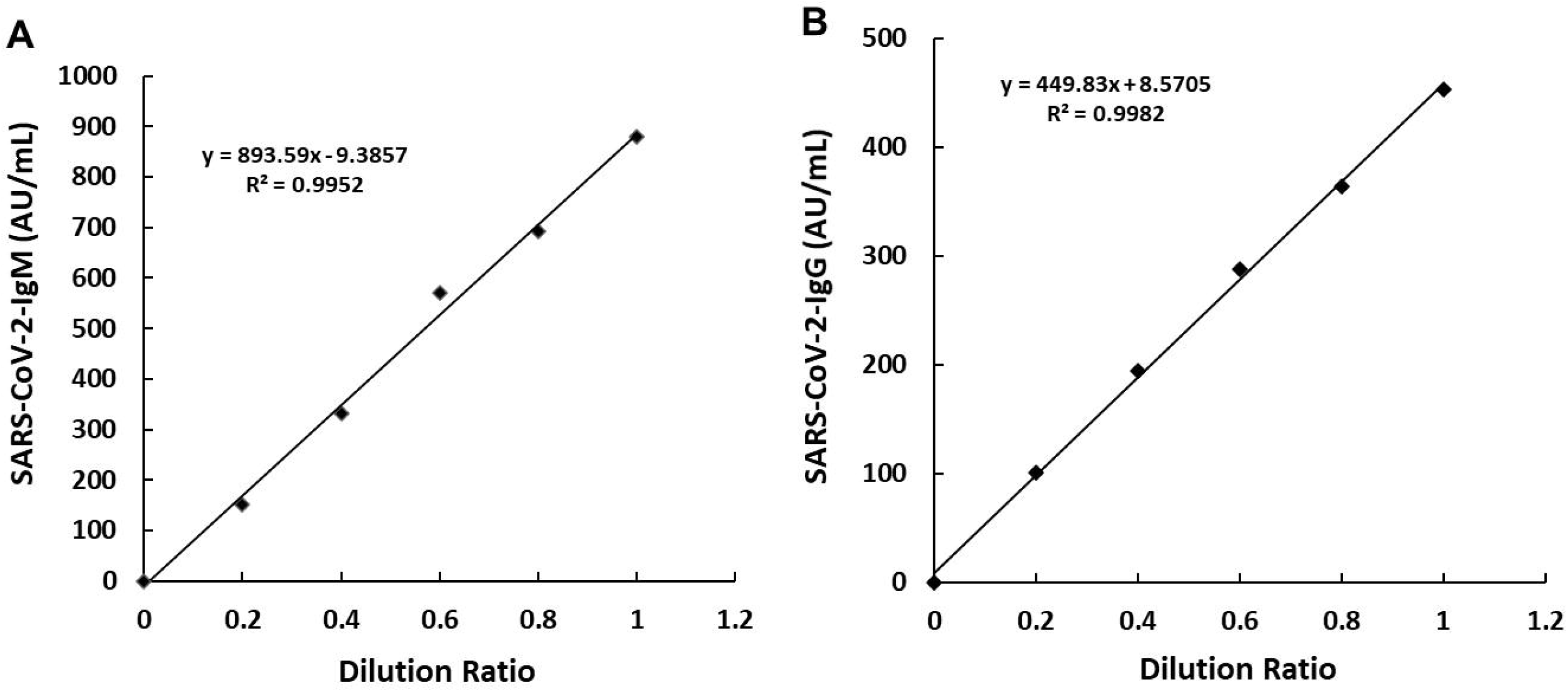
Linearity of SARS-CoV-2 IgM (A) and SARS-CoV-2 IgG (B)

### CLINICAL SPECIFICITY

Samples from both hospitalized patients (972 subjects) and normal population (586 subjects) were used to assess the clinical specificity of the assay (Table 3). SARS-CoV-2 IgM shows the clinical specificity of 97.33% for hospitalized patients and of 99.49% for normal population. SARS-CoV-2 IgG shows the clinical specificity of 97.43% for hospitalized patients and of 99.15% for normal population.

**Table 3:**
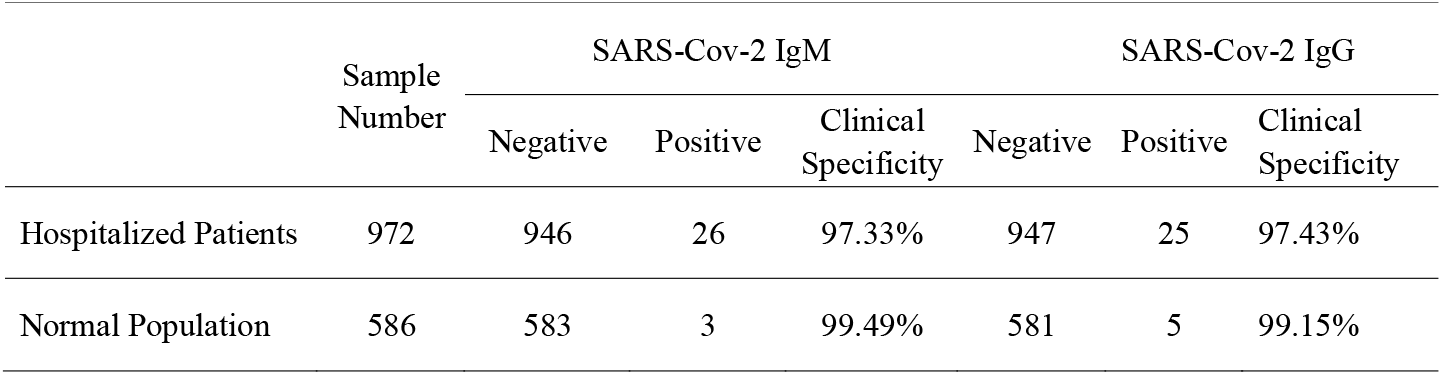
Clinical specificity of SARS-CoV-2 IgM and SARS-CoV-2 IgG

### CLINICAL SENSITIVITY

Samples from both confirmed SARS-Cov-2 infection with RT-PCR (503 patients) and the suspected SARS-Cov-2 infection (52 patients) were used to evaluate the clinical sensitivity of the assays (Table 4). SARS-CoV-2 IgM and IgG show a clinical sensitivity of 85.88% and 96.62% respectively for the patients confirmed SARS-Cov-2 infection with RT-PCR. The study recruited 52 patients whom had been highly suspected for SARS-CoV-2 infection based on typical epidemiological history, clinical symptoms and featured CT image, but the viral nucleic acid remained negative after more than 3 attempts of detection with RT-PCR. When measuring the antibodies with SARS-CoV-2 IgM and IgG immunoassays, 38 of them show the positive result for SARS-CoV-2 IgM, giving a detection rate of 73.08%, and 45 of them are SARS-CoV-2 IgG positive, giving a detection rate of 86.54%.

**Table 4:** Clinical sensitivity of SARS-CoV-2 IgM and SARS-CoV-2 IgG

|  | Sample<br>Number | SARS-Cov-2 IgM |  |  | SARS-Cov-2 IgG |  |  |
| --- | --- | --- | --- | --- | --- | --- | --- |
|  |  | Negative | Positive | Clinical<br>Sensitivity | Negative | Positive | Clinical<br>Sensitivity |
| Confirmed SARS-Cov-2<br>infection (RT-PCR positive) | 503 | 71 | 432 | 85.88% | 17 | 486 | 96.62% |
| Suspected SARS-Cov-2<br>infection (RT-PCR negative) | 52 | 14 | 38 | 73.08% | 7 | 45 | 86.54% |

## Discussion

During the outbreak of COVID-19, we took the lead in developing automatic SARS-CoV-2 IgM and IgG immunoassays, and performed a multi-center evaluation with a large population across 10 hospitals in the Wuhan epidemic area and hospitals in other provinces in China. The assays are based on chemiluminescence detection technology, and run on an automatic immune analyzer. They automatically measure the specific IgM and IgG antibodies to SARS-CoV-2 using peripheral blood, and the time to the first result is less than 30 minutes at a speed of more than 150 tests per hour, which would allow for the screening of COVID-19 in large populations. SARS-CoV-2 IgM and IgG immunoassays demonstrated satisfied assay range and precision with a CV of less than 4.45%, meeting the standard for identifying SARS-CoV-2 infection in clinical laboratories.

As COVID-19 is a virulent infectious disease, inactivation of the specimen at 56°C for 30 minutes is necessary to avoid the potential risk of infecting laboratory technicians. To verify whether inactivation affects the detection for the SARS-CoV-2 antibodies, specimens from 55 subjects were studied, including 15 normal subjects and 40 SARS-CoV-2 IgM and/or IgG positive patients. No significant influence on the levels of IgM and IgG antibodies to SARS-CoV-2 was found, and no false positive or false negative observed, indicating that inactivation of specimen does not affect the measurement of the assays. We therefore recommend a routine specimen inactivation to ensure the safety of the contact people.

Detection of SARS-CoV-2 viral nucleic acid is a direct evidence of viral infection, thus RT-PCR has been thought to be the gold standard for etiological diagnosis of COVID-19. However, due to the typical un-productive cough symptom of COVID-19, the occurrence of SARS-CoV-2 in the upper respiratory track is relatively low, leading to challenges in collecting specimens through nasopharyngeal swabs. Furthermore, the viral RNA is not stable and degrades easily after specimen inactivation, causing false negatives in nucleic acid tests. There are many cases in Wuhan as well as in other provinces in China where the nucleic acid test of a recovered patient turned positive after being discharged from hospital for some days, increasing the risk of infecting others and missing the opportunity for proper treatment. Very recently on February 26, a patient who was considered cured after testing negative on two consecutive nucleic tests, was discharged from a hospital in Wuhan. Five days later on March 2, the patient was hospitalized again and died of COVID-19 (unpublished data). The reason why nucleic acid tests are considered gold standard is because a diagnosis can be confirmed once it is positive. However, the inverse is not true, as a negative result does not rule out viral infection. SARS-CoV-2 are novel coronaviruses, and very little is known about the status of viral reproduction in the body. For a certain period after the infection the virus may incubate in the body and stop excretion after medical intervention, leading to a false negative on a nucleic acid test. It is critical for an alternative testing method to be developed to compensate for this limitation of nucleic acid testing.

Antibodies are proteins produced by the immune system after being stimulated by viral pathogens, and usually very little viral antigen can stimulate the body to produce enough antibodies that can be detected by sensitive immunoassays. The generation and detection of antibodies to viruses are usually not affected by the status of virus in the body, even during the dormant phase of viruses. In addition, as peripheral blood is used for antibody detection, sample collection is easily controlled than nasopharyngeal swabs and is not influenced by a sample collector’s operational deviation, ensuring a better repeatability of the test results. It also does not impose higher biological risk for sample collectors.

This study evaluated SARS-CoV-2 IgM and IgG immunoassays for their clinical specificity using serum from 586 healthy individuals, and the specificity of both the IgM and IgG assays exceeded 99%. When evaluated with 972 serum samples from hospitalized patients diagnosed with diseases other than COVID-19, the two assays demonstrated clinical specificity of greater than 97%. Due to interference on immunoassays, the specificity based on inpatient samples is usually lower than that based on healthy population, especially the immunoassays for markers of infectious diseases. Many factors could be involved in affecting the specificity of SARS-CoV-2 IgM and IgG immunoassays, including autoimmune diseases, tumors, infections with other coronaviruses and flu viruses. Further studies are needed to verify the sources of interference and cross-reactivity to the assay.

The outbreak of the COVID-19 in Wuhan allowed us to recruit a large population of infected patients to evaluate the sensitivity of SARS-CoV-2 IgM and IgG immunoassays. Serum from 503 patients confirmed by RT-PCR nucleic acid tests have been measured for SARS-CoV-2 IgM and IgG concentration, and the clinical sensitivity exceeded 85% and 96% for SARS-CoV-2 IgM and IgG respectively. It should be noted that nucleic acid tests had been performed at the early onset of the symptoms of COVID-19, and were often repeated several times for a positive confirmation. The measurement of serum antibodies for SARS-CoV-2 IgM and IgG took place at relatively later stage, starting from February 17 when the assays were available. Therefore, the levels of serum IgM in some patients could have started to decline to a level that could not be detected by the assay. We speculate that this could be the major reason why the clinical sensitivity (detection rate) of IgM is lower than that of IgG antibodies.

The reason why newly confirmed COVID-19 cases in Hubei province increased drastically to 13,332 people in one-day on February 12, was because of the release of a new set of guidelines for the diagnosis and treatment of COVID-19 (version 4) *(15)*, in which the etiological history, typical clinical symptoms and the featured chest CT scanning were included for a diagnosis of COVID-19, despite a negative result of RT-PCR. The majority of the accumulated “suspected COVID-19 cases” were actually the SARS-CoV-2 infection, but due to the shortage and the considerable false negative results of nucleic acid testing, the cases could not be confirmed by laboratory diagnosis. With the development and clinical application of the immunoassays, the detection rate for COVID-19 cases had significantly increased, prompting the release of a new set of guidelines to the diagnose and treatment of COVID-19 (version 7) *(16)*, which included antibody detection as one of the laboratory diagnostic criteria for confirming COVID-19. We have examined 52 suspected cases for COVID-19 whose nucleic acid tests had repeatedly returned as negative, and found that 38 of them were SARS-CoV-2 IgM positive and 45 of them were IgG positive, indicating examination of the specific SARS-CoV-2 antibodies can efficiently compensate for the false negative limitations of nucleic acid testing.

Study on the sensitivity for convalescent samples is critical for evaluating the performance of the assays for infectious diseases, as this represents the ability for early detecting an infection, especially the diseases like COVID-19. Due to the limited time after developing SARS-CoV-2 IgM and IgG assays and the access limitation for patient information, we have only collected some preliminary data on the sensitivity for convalescent samples. On day 3 after the onset of the symptom, we observed two patients, one of whom with IgG concentration of 144 AU/mL and IgM of 20.34 AU/mL (cutoff = 10 AU/mL), and the other patient with IgG of 156.91 AU/mL and IgM of 9.79 AU/mL (close to the cutoff of 10 AU/mL). However, on day 5, we recruited one patient with negative for both IgM and IgG. From day 6, most of the patients were positive for IgM and/or IgG. Therefore, we speculated that the serum antibodies for both IgM and IgG could be detectable with the assays at around day 6 after the onset of the symptom. We saw the trend of IgM increased from day 6, peaked at around day 20, and then kept decreasing till undetectable on about day 35, while there no significant trend observed for IgG at the small period of time window from day 6 to day 35. It should be noted that those preliminary data need to be further investigated with more samples, especially the samples from early onset patients.

In conclusion, we have developed fully automated immunoassays for the detection of SARS-CoV-2 IgM and IgG antibodies in human serum using an automatic immune analyzer. Using specimens of peripheral blood, the immunoassays solved the sample collection challenges of nucleic acid testing. Thanks to the full automation of the immunoassays, sophisticated operational training and stringent laboratory settings are not required, as is the case for nucleic acid testing. The high throughput of SARS-CoV-2 IgM and IgG assays allows for the mass screening for COVID-19. The high clinical specificity and sensitivity of the automated SARS-CoV-2 IgM and IgG immunoassays add great value to nucleic acid testing in fighting against COVID-19.

## Data Availability

The data that support the findings of this study are available from the corresponding author on reasonable request.

## Research Funding

B.F. Liu, Fundamental Research Funds for the Central Universities (2020kfyXGYJ103).

## Nonstandard abbreviations

SARS-CoV-2: severe acute respiratory syndrome coronavirus 2
COVID-19: novel coronavirus disease
CLIA: chemiluminescent immunoassay
RT-PCR: Real Time-PCR

